# Computerized physical exercise improves the functional architecture of the brain in patients with Parkinson’s Disease: a network science resting-state EEG study

**DOI:** 10.1101/2020.10.21.20209502

**Authors:** Vasileios Rafail Xefteris, Charis Styliadis, Alexandra Anagnostopoulou, Panagiotis Kartsidis, Evangelos Paraskevopoulos, Manousos Klados, Vasiliki Zilidou, Maria Karagianni, Panagiotis D. Bamidis

**Author notes:** **Corresponding Author:** Dr. Charis Styliadis, School of Medicine, Faculty of Health Sciences, Aristotle University of Thessaloniki, Thessaloniki, Greece. Aristotle University of Thessaloniki, University Campus, P.C. 54124, Thessaloniki, Greece.

## Abstract

Physical exercise is an effective non-pharmaceutical treatment for Parkinson’s disease (PD) symptoms, both motor and non-motor. Despite the numerous reports on the neuroplastic role of physical exercise in patients with PD (PwPD), its effects have not been thoroughly explored via brain network science, which can provide a coherent framework for understanding brain functioning. We used resting-state EEG data to investigate the functional connectivity changes of the brain’s intrinsic cortical networks due to physical exercise. The brain activity of 14 PwPD before and after a ten-week protocol of computerized physical training was statistically compared to quantify changes in directed functional connectivity in conjunction with psychometric and somatometric assessments. PwPD showed a significant reorganization of the post-training brain network along with increases in their physical capacity. Specifically, our results revealed significant adjustments in clustering, increased characteristic path length, and decreased global efficiency, in correlation to the improved physical capacity. Our results go beyond previous findings by indicating a transition to a reparative network architecture of enhanced connectivity. We present a meaningful relationship between network characteristics and motor execution capacity which support the use of motor treatment in tandem with medication. This trial is registered with ClinicalTrials.gov Identifier NCT04426903.

**Impact Statement:** The effects of physical training (PT) on the neuroplasticity attributes of patients with Parkinson’s Disease (PwPD) have been well documented via neurophysiological evaluations. However, there is a knowledge gap on the role of training-induced neuroplasticity in whole-brain network organization. We investigated the PT effects on the brain network organization of 14 PwPD, using EEG and network indices coupled with psychosomatometric tests. We report evidence of reparative functional reorganization of the brain with more balanced integration and segregation abilities, in correlation to improved motor performance. The PD brain can repair and reestablish a better level of motor execution and control due to computer-empowered physical stimulation.

## Introduction

Neuroplasticity is the brain’s ability to adjust and reorganize itself across an organism’s lifespan in both health and disease [1]. Patients with Parkinson’s Disease (PwPD), in contrast to healthy old adults, experience loss of dopaminergic brain cells of the substantia nigra projecting to the striatum [2]. Dopamine influences synaptic plasticity [3], and thus, dopaminergic neural loss in PD leads to an impaired neuroplastic ability [4].

PD progression is characterized by brain atrophy, mainly in anterior brain regions but also extending to posterior ones [5], [6], as well as disconnections between brain regions [7]–[10]. Consequently, PD has been described as a disconnection syndrome with relevance to cognition, perception, and other neuropsychological domains [11]. Clear implications of this notion emerge from the study of the brain’s intrinsic resting-state networks (RSNs). For instance, altered connectivity of the default mode network (DMN) can have a detrimental effect on the cognitive capacity of PwPD [9], [10], [12], [13], while temporal-occipital disconnections between the visual (VIS) network and DMN have been correlated with visuospatial impairment [9], [13]. Disruptions in the interconnections of the DMN and the frontoparietal network (FPN) can affect both executive functioning and cognition [12], while disturbances in the sensorimotor network and somatomotor (SMN) one have been found to relate to motor symptoms of PD [7], [9], [14].

A healthy human brain organizes itself in what is called a small-world (SW) network. SW organization features balanced local (local clustering of connections) and global (long-range connections) efficiency; both deemed essential for healthy brain function [15]. The brain network architecture of PwPD shows disruption of SW architecture towards a more random brain organization [9], [16]. This disruption is reported even at the very early stages of the disease, meaning lower segregation (local clustering), but preserved integration (average path length) [9], [16]. Along with the disease’s progression, the brain’s functional architecture further deviates from the SW organization as it loses both local and global efficiency in correlation to PD-related cognitive decline [16], [17].

Though one would expect that network abnormalities would limit plasticity, the PD brain can still feature neuroplastic capabilities, at least as compensatory mechanisms [18], [19]. For instance, PD-related motor deficits only arise after approximately 80% depletion of striatal dopamine [20], and thus, the underlying compensatory mechanisms delay the emergence of clinical symptoms in PD [18]. This compensatory reorganization is more dominant in the motor tracts, suggesting selective neurodegeneration [19].

From a therapeutic perspective, only pharmaceutical symptomatic treatments are available, often having potential side effects [21]. In contrast, neuroscientific evidence on brain-derived neurotrophic factor (BDNF) changes [22] or volumetric changes [23] in PwPD have shown promise for effective physical training (PT) protocols, which can trigger neuroplasticity and neuroprotection [24]. For instance, non-computerized PT can benefit muscular strength, mobility, and postural disabilities of PwPD [25], improving their confidence [26] and quality of life [27]. Computer-based training has shown to be profitable for healthy elders [28] and elderly with neurodegenerative disorders, e.g., mild cognitive impairment (MCI) [29]. Despite the rare exploitation of computerized PT by means of (serious) game-based training, there are promising findings such as improved balance [30], rate of falls, and confidence in PwPD [31].

Given the network disruption in PD [32], [33], invegistations into neuronal network reorganization due to PT may be helpful to bridge the gap between the mechanisms underlying the effectiveness of non-pharmateutical interventions and clinical presentations in PD. To our knowledge, only a few studies have examined functional connectivity changes following a non-computerized PT program [34]–[36] but these focused on the motor circuitry. Evidence in regard to computer-assisted PT for PwPD, though more scarce, has shown whole-brain network connectivity changes, and suggests that treadmill training with virtual reality can offer better results in PwPD (i.e. greater connectivity within the sensorimotor network and the cerebellar network in association with improved walking performance) [37].

Here, we aim to narrow the knowledge gap in PD literature regarding the reorganization of the whole brain architecture and the relevant changes in network characteristics as a result of PT while at the same time offer innovative means and tools to facilitate such training. We hypothesized that our computerized PT scheme, combining aerobic, flexibility, strength, and balance exercises, could trigger the reorganization of the brain network in a reparative manner, namely towards a network architecture of SW properties. To this aim, we used eyes-closed resting-state electroencephalographic (EEG) recordings before and after the 10-week long WebFitForAll [28] intervention. Source analysis was performed using low resolution electromagnetic tomography (LORETA) [38], and directed functional connectivity was estimated via phase transfer entropy (PTE) [39] in the 1-30 Hz frequency band. An FDR corrected t-test statistical comparison was performed to identify the significant connectivity changes (post-vs. pre-intervention). We also computed selected graph-theoretical parameters that describe integration, segregation, and hierarchy to characterize plasticity in the PD brain and investigate how the training-induced neuroplasticity affects the state of the PD network.

## Results

### Psychometric and somatometric results

Psychosomatometric comparisons between the pre- and post-intervention scores were performed using non-parametric Wilcoxon tests and paired t-tests. We found statistically significant increases in the scores of the Berg Balance Scale (BBS) [40], the Short Physical Performance Battery (SPPB) [41] balance and total score, the Community Balance & Mobility Scale (CB&M) [42], the Fullerton Senior Fitness Test (SFT) [43] Arm curl and 2-min Step test, the Tinetti Performance Oriented Mobility Assessment (POMA) [44], Gait and Total scores and decrease of the time of the 10 Meter Walk Test [45] on both self-selected and fast velocity and SFT 8-foot Up-and-Go test. No statistically significant differences were found in the psychometric evaluation tests (i.e., Mini-Mental State of Examination (MMSE) [46], Montreal Cognitive Assessment (MoCA) [47], and Parkinson’s Disease Questionnaire (PDQ-8) [48]).

### PTE results

Statistical comparisons of the PTE matrices of the post vs. pre-intervention EEG data indicate the reorganization of the PD resting-state network (45 nodes, *p < 0*.*05*, FDR corrected, 10000 permutations) (Figure 1). Specifically, the cortical reorganization is characterized by the enhanced connections between: (i) middle and inferior occipital nodes and parahippocampal, and middle and superior temporal nodes of the left hemisphere, and inferior frontal, culmen, and fusiform nodes of the right hemisphere, and (ii) right (middle and superior) frontal nodes and inferior frontal, parahippocampal, middle occipital, and middle and superior temporal nodes of the left hemisphere. In respect to the core resting-state networks, as defined by Yeo et al.[49], we have noted increased connectivity within the VIS, and between the VIS and each of the DMN, the dorsal attention network (DAN), the ventral attention network (VAN), SMN, and limbic (LIM) networks, the DMN and the SMN, as well as the FPN and the LIM, the VIS and the SMN. Figure 1 illustrates the statistically significant post to pre-intervention differences of the brain networks. For illustrative purposes, we also provide a circular graph of the brain connectivity network. Table 2 summarizes the connections between the core resting-state networks.

**Table 1.** Significant post vs. pre differences on somatometric tests and the post-pre mean difference. The sign of the mean difference denotes the direction (effect) of the changes. Results were considered significant for *p < 0*.*05*.

| Somatometric tests | p-value | t-value | Mean difference |
| --- | --- | --- | --- |
| BBS | 0.019 | 2.326 | 1.71429 |
| SPPB Balance | 0.028 | 2.482 | 0.42857 |
| SPPB Total | 0.026 | 2.511 | 0.57143 |
| CB&M | 0.003 | 3.215 | 6.85714 |
| SFT Arm Curl | 0.028 | 2.349 | 2.357 |
| SFT 2-min Step | 0.015 | 1.172 | 7.714 |
| SFT Up-and-Go | 0.040 | -2.263 | -1.25714 |
| 10-meter time Self | 0.022 | -2.263 | -0.51500 |
| 10-meter time Fast | 0.011 | -3.126 | -0.27786 |
| POMA Gait | 0.033 | 2.110 | 0.71429 |
| POMA Total | 0.043 | 2.008 | 0.92857 |

**Table 2.**
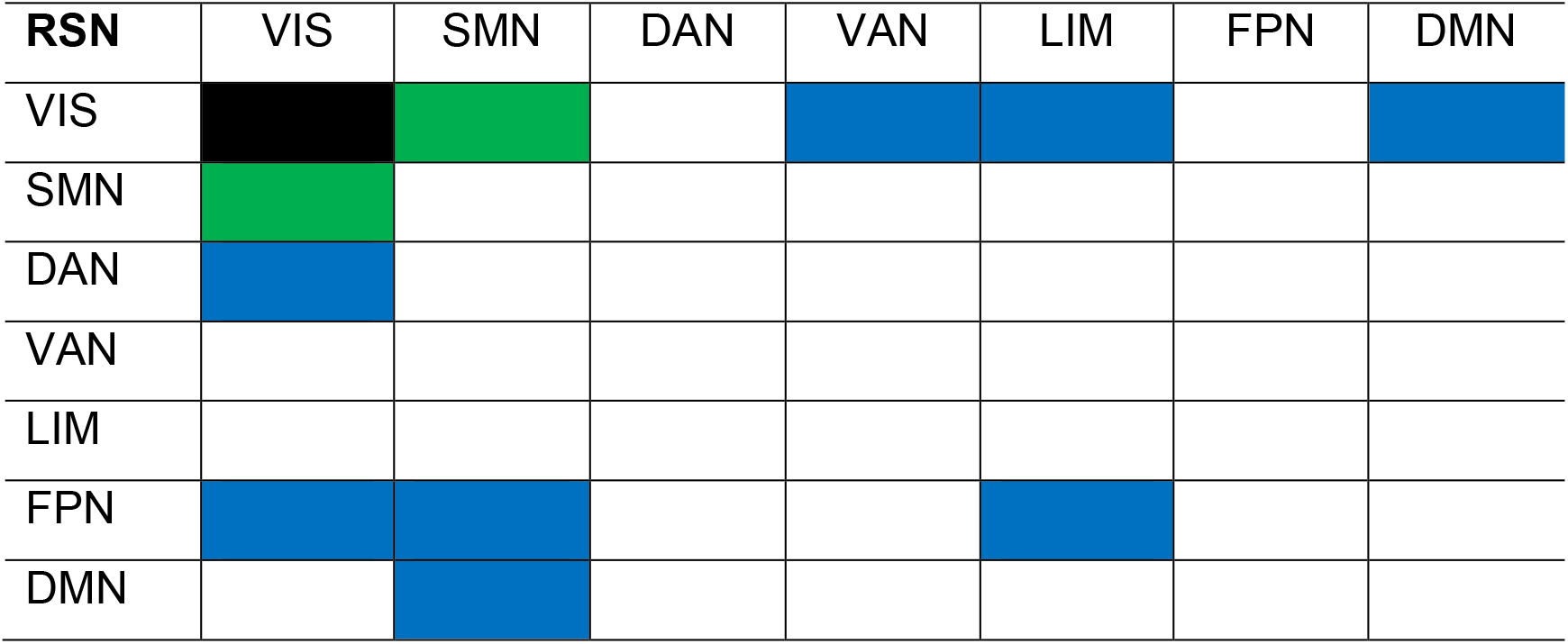
Significant increase in connectivity of RSNs as defined by Yeo et al [49]. The black color signifies the within-network connectivity, the green color the bilateral connectivity between-networks, and the blue color the unilateral connectivity between-networks from the RSN in the first column to the RSN in the first row.

**Figure 1.**
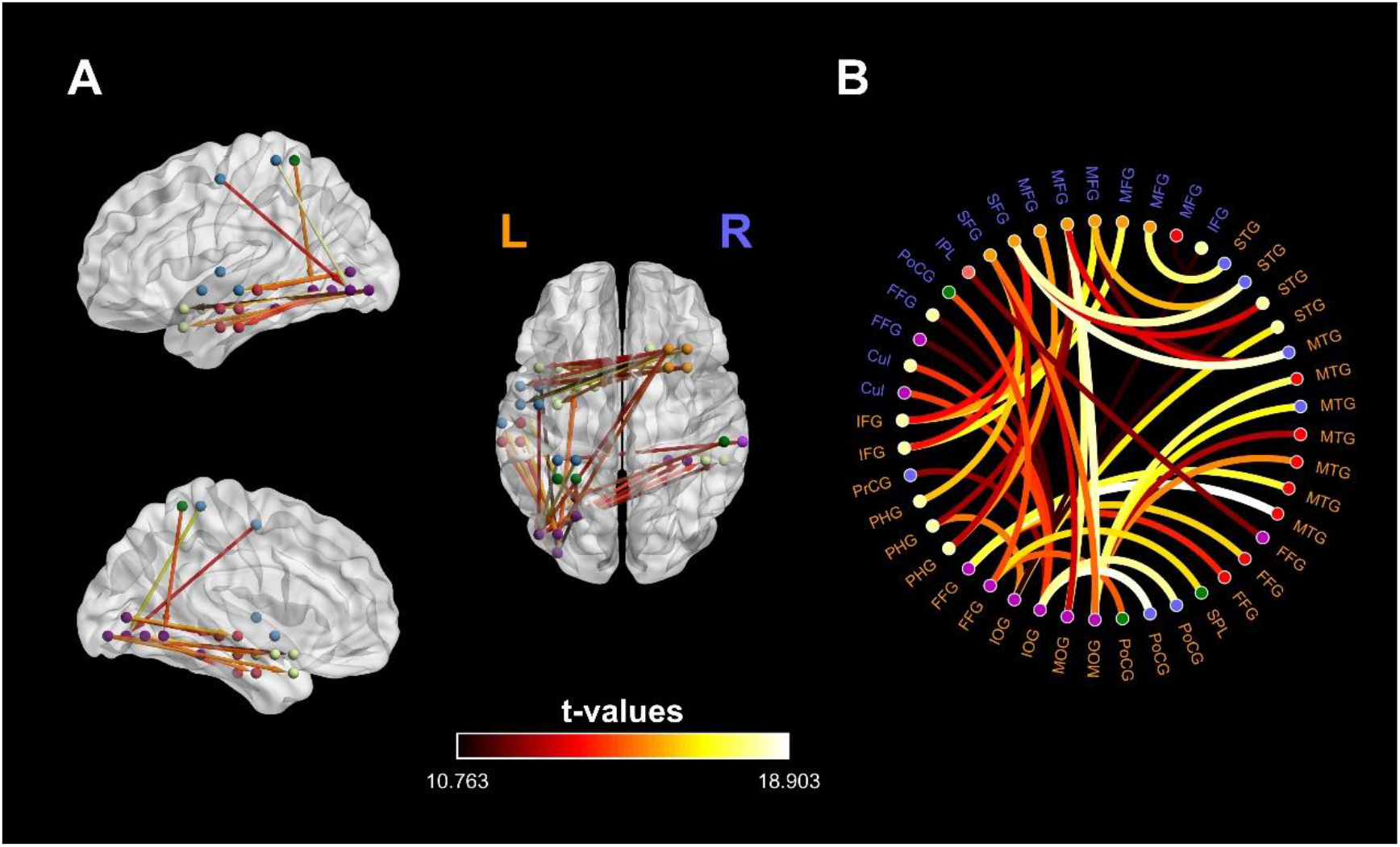
Significant differences in cortical connectivity between post- and pre-intervention networks (orange denotes the left hemisphere and blue the right). Node colors correspond to distinct intrinsic resting-state networks [49] (Purple-Visual, Blue-Somatomotor, Green-Dorsal Attention, Violet-Ventral Attention, Cream-Limbic, Orange-Frontoparietal, Red Default Mode). **A:** Information direction is depicted through line arrows, and the color scale represents t-values. The visualized networks are significant at a level of p<0.05, FDR corrected. **B:** Circular graph depicting the cortical reorganization in the PD brain. The cortical reorganization is characterized by the emergence of a direct connection between the left inferior occipital gyrus, the left middle occipital gyrus, the left middle temporal gyrus, the left superior temporal gyrus, the right middle frontal gyrus, and the fusiform gyrus.

### Graph Measures

Graph theory analysis included measures of integration (Global Efficiency (GE), Characteristic Path Length (CPL)), segregation (Cluster Coefficient (CC), Transitivity (TS), Local Efficiency (LE), Modularity (Q), and node centrality (Betweenness Centrality (BC), Degree Centrality (DC)). Our graph-theoretical analysis, using each subject’s adjacency matrix, revealed statistically significant differences in the GE, CPL, and TS (Table 3). We report a decrease in the GE and TS and an increase in the CPL due to our intervention. We found statistically significant increases and decreases in the local CC. The increase of CC was observed in 210 nodes, including bilateral frontal, temporal, parietal, and lingual lobes, and the right occipital lobe. The remaining 653 nodes showed reduced CC after the intervention. Figure 2A depicts the nodes with increased CC, and Figure 2B those with decreased CC. LE, BC, DC, and Q did not show any statistically significant changes.

**Table 3.**
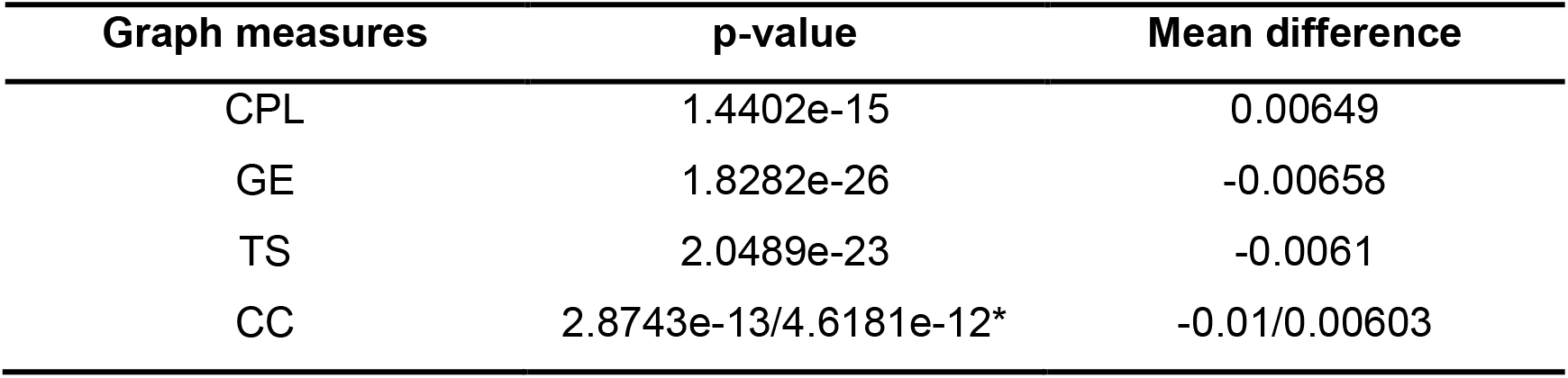
Significant post-pre changes in global (Global efficiency, Transitivity, Characteristic Path Length) and local (Local Clustering Coefficient) graph measures, along with their mean differences. The sign of the mean difference denotes the direction (effect) of the changes. Results were considered significant for *p < 0*.*05*. *For the local graph measures, the p-value reported is the mean value of the nodes showing the significant increase and decrease, respectively.

**Figure 2.**
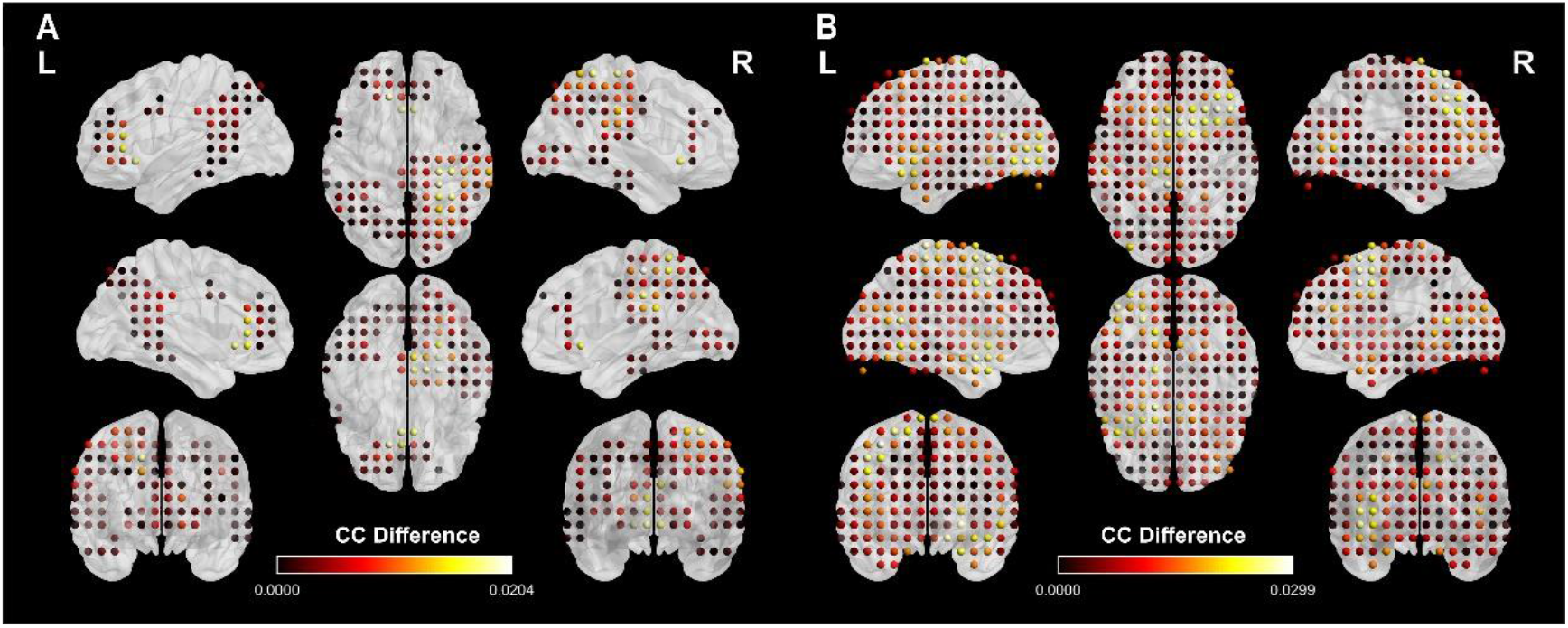
Whole-brain map of the statistically significant changes of the CC of the 863 nodes. **A:** Increased CC in 210 nodes of bilateral frontal, temporal, parietal, and lingual lobes and right occipital lobe. **B:** Decreased CC in 653 nodes of bilateral frontal, lingual, occipital, parietal, and temporal lobes.The color map indicates the significance of the CC difference.

### Multiple Regression

We tested for relations between network science indices and physical performance scores through linear regression analysis and found a statistically significant correlation between changes of SPPB total scores and graph theory indices of GE, CPL, and TS. We report that SPPB total score can predict GE (p= 0.011, a = −0.092, b =0), CPL (p = 0.007, a = 0.095, b = 0) and TS (p = 0.012, a = −0.092, b =0) changes. This means that higher scores in the SPPB test can potentially hint to the increase in CPL and decreases in GE and TS.

## Discussion

Our findings indicate that a ten-week intervention of computerized physical training, in patients with PD, facilitates neuroplasticity, which consequently induces a beneficial cortical reorganization. Specifically, our results reveal that: (i) short-term PT influences positively the physical performance of PwPD, (ii) the PD brain can utilize neuroplasticity to reorganize itself in a reparative manner, (iii) reparative neuroplasticity may lead to a more flexible functional network, and (iv) the reparative reorganization correlates to the improvement of motor execution and control.

### Short-term training influences physical performance

The statistically significant differences in mobility, static and dynamic balance, muscular strength and endurance, functional mobility, and gait speed and control, and reduced risk of falls (Table 1) are in line with previously reported findings [27], [50]. Here, we report an improved motor performance in PwPD as an outcome of PT. It must be noted that our protocol targets multiple domains (i.e., aerobic, resistance, balance, and flexibility exercises), while most studies investigating the effects of PT on the somatic capacity of PwPD have focused mainly on a specific domain, i.e., strength [27] or aerobic [51] and, less often, on a combination of two or more domains [50].

### Cortical reorganization of the PD brain network

Our results indicate the emergence of a reorganized brain network of enhanced connectivity, triggered by our computerized PT intervention, which involves connections mainly within the VIS and between the DMN and the VIS, and connections of the FPN with the LIM, VIS, and the SMN (Figure 1, Table 2). The enhanced connectivity within and between the RSNs indicates that PwPD present capacity for neuroplasticity and to functionally reorganize their brains. Our findings go beyond previous studies that examine the results of PT focusing only on the connectivity of the primary motor cortex [34]–[36]. By exploiting the innovations brought about by the use of a fully computerized program for PT and utilizing whole-head analysis, we report the arise of effective connections, thus provide further insight into the neuroplastic attributes of PwPD due to computerized PT.

The increased connectivity between the DMN and VIS hints towards the neuroprotective role of physical exercise in PD. The cognitive capacity of PwPD has been previously reported to positively correlate with the connectivity of the DMN and VIS, meaning reduced connectivity leads to reduced cognitive capacity, with the occipital gyrus having a key role [52], [53]. The patients’ cognitive deterioration (i.e., in MCI) is accompanied by a loss of connections in the DMN, an effect evident from the early stages of the disease [54], meaning that cognitive decline in PD can be associated with disrupted connections in the DMN even in cognitively unimpaired PwPD [10]. As PD progresses, the occipital lobe exhibits hypoconnectivity with the temporal and frontal lobe [55], with changes in the visual system manifesting even before visual symptoms become clinically evident [56]. Additionally, the disrupted connectivity between the frontal and occipital cortex might also relate to typical non-motor symptomatology of PD, such as executive dysfunction, attention problems, or lower performance in visuospatial tasks [13]. We report increased connectivity between the DMN and VIS (Figure 1, Table 2), with the direction of information, as reflected by the PTE, indicating that the left occipital gyrus modulates the activity of the left middle temporal gyrus of the DMN. These shifts in the connectivity of PwPD indicate the formation of a network that points to the training’s reparative effect.

Regarding FPN’s connections to other RSNs, the FPN seems to work here as a hub modulating LIM, VIS, and the SMN (Table 2), in line with its previously reported role as an adjustable hub among other networks for the flexible coordination of cognitive control [57]. The increased information flow from the FPN towards the LIM and the VIS networks could be interpreted as a mechanism of cognitive control and reorientation. Nevertheless, this assumption is not supported by our psychometric assessments. The modulation of the SMN from the FPN, combined with the statistically significant differences of the somatomotor assessments’ scores, could index an information pathway offering improved motor control and execution.

#### Global and local graph indexes reveal a transition towards a reparative PD network organization

Considering that the PD brain network features an abnormal structure towards randomness [16], we suggest that the reorganization of PD brain circuitry, due to PT, is characterized by a transition from a random-alike network towards a network architecture of SW-alike properties. Even though the reported decreased GE and increased CPL might indicate a step towards a less efficient network organization, this contradictory result could be explained as follows. Networks can be demonstrated inside a framework of regular, random, and complex networks. Regular networks appear to have high local and low global connectivity, while random networks show the contrary design with low local and high global connections [58]. The optimal function of human brain network architecture balances between integration and segregation by incorporating elements of regular and random networks to create a more complex architecture, the so-called SW organization [58]–[60]. Here, decreased GE and increased CPL, coupled with the fact that the PD brain network has a random-alike structure, indicates excessive network integration in PwPD. This is further supported by evidence that as cognitive impairment becomes more severe, the network of PwPD shows higher GE and lower CPL, indicating more randomness [61]. Thus, our findings of lower GE and higher CPL could index the triggering of reparation neuroplasticity.

It is plausible that the transition’s attributes would at some point (e.g. due to more intense and/or longer duration of the training) reflect a SW network topology (Figure 3). Here, the SW-alike attributes are present not only on a local level as well. Changes of the CC (Figure 2) indicate a more robust functional clustering organization, further supporting our suggestion of a reparative brain network architecture. Our post-training network’s robustness and segregation can provide neuroprotection in PwPD [16], but the efficacy of such a mechanism depends on the balance between integration and segregation, featuring SW network architecture [58], [60].

**Figure 3.**
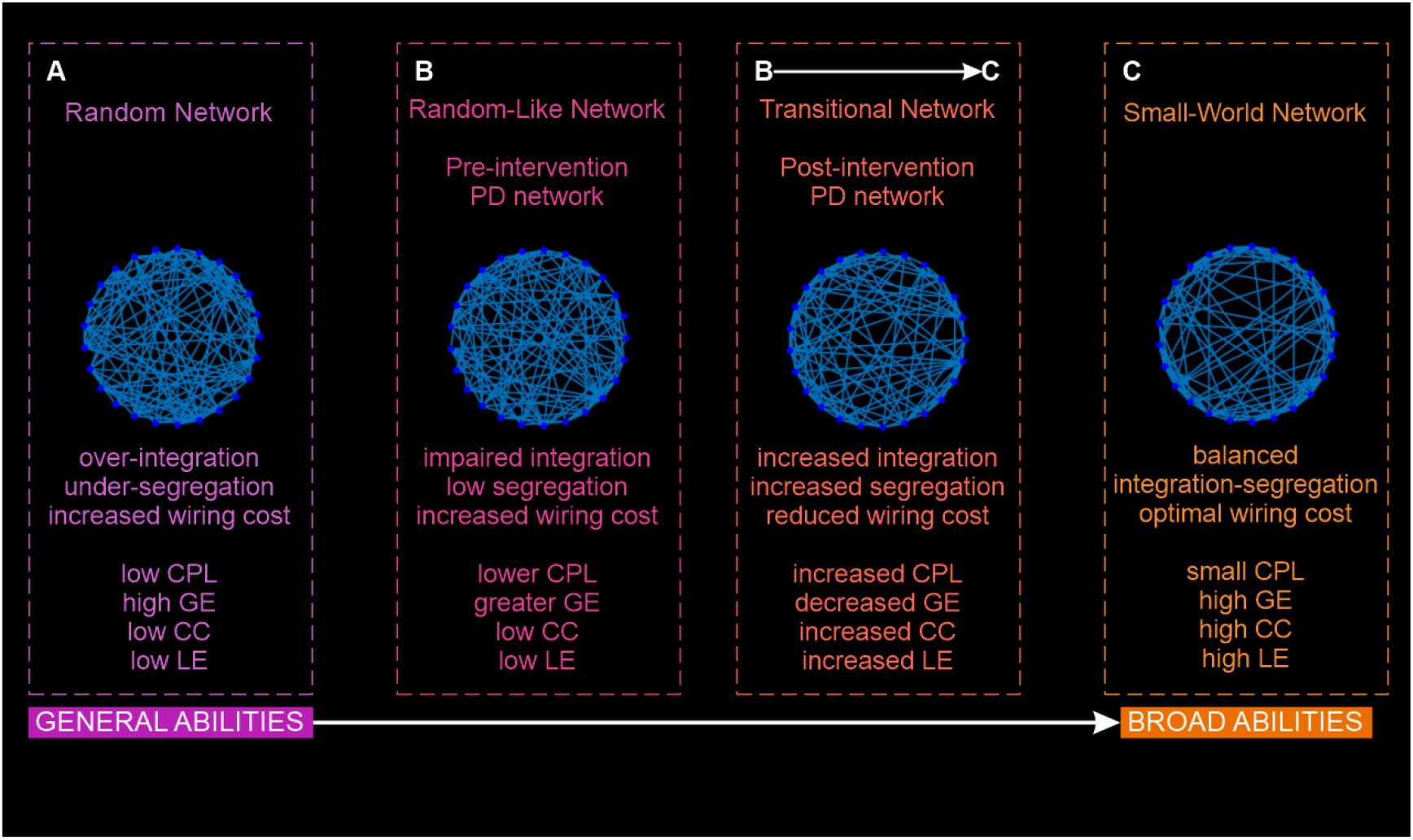
Illustration of our theoretical proposal. High integration (high GE) is characteristic of random networks (A), while regular networks show higher segregation (high LE). The optimal brain network organization is that of a SW network (C) balancing between high integration and segregation achieving the optimal wiring cost [15]. As reported by Dubbelink et. al [16] PD brain network organization, compared to controls, resembles that of random networks having less SW network characteristics (pre-training network, B). Changes in connectivity and graph-theory characteristics, as an outcome of the reparation neuroplasticity in the PD brain, characterize our post-training network (B → C) as a transitional network from a random-alike network (B) towards a SW-alike organization (C), meaning more balanced integration and segregation resulting in a restored brain organization for our PwPD.

#### Whole-brain network reorganization improves motor execution and control

Our multiple linear regression analysis revealed that SPPB total score correlates negatively with the post to pre differences of GE and TS, and positively with CPL post to pre changes. The higher total score of SPPB means reduced GE and TS and increased CPL. There is a relationship between better motor performance and reorganization of brain network connectivity patterns towards a more efficient architecture; better motor execution predicts a transition towards a reparative functional architecture, balancing both random and regular network characteristics. The improvements in gait, mobility, and balance, as aspects of the SPPB test [41], meaning improved preparation and control of movement, is correlated to a more efficient brain organization with more SW-alike characteristics. This correlation further supports our suggestion of a PT-induced network organization that can prepare for, and control motor tasks.

Motor severity in PD is correlated with the mean functional connectivity strength [9], meaning that disconnections observed in PD brain circuitry are linked to motor symptoms. The loss of integration in PD’s network as the disease progresses is associated with motor disorientation and motor symptoms severity [16]. Here, we found not only a balance of excessive integration (increased CPL and decreased GE) as a PT result, but we report that this balance is connected to improved motor performance. Thus, we can imply that these indexes hold promise as surrogate markers of motor symptoms severity in PD.

#### Future directions and limitations

Our relatively small sample size (14 PwPD) and the lack of an age-matched control group (healthy old adults or passive PwPD) may have limited the interpretation of our findings. Accordingly, the effects of our intervention could have been classified concerning different PD motor subtypes (e.g. tremor-dominant or postural instability gait difficulty) or along the continuum of cognitive capacity (e.g. PD-MCI). The correlation of the improved motor execution with a reparative brain organization is in support of the beneficial contribution of PT to disease modification and points towards the use of physical activity as a supplement to pharmacotherapy. Future studies could be targeted in disentangling the elements of neuroplasticity induced by physical (and/or cognitive) training and identify their contribution to disease modification. Follow-up studies are essential to address the stability of the reported findings. Though the modulation of the DMN from the occipital gyrus could mean improved cognition in PwPD, this was not evident in our study. Future reports could provide more insight into the possible relation of PT-induced reparative neuroplastic and cognitive capacities in PD.

#### Conclusion

Our study provides emphasizes the need to introduce physical exercise in the daily activities of PwPD. In contrast to previous studies that focused on the primary motor area, the focus here is on the whole-brain network level. PT facilitates neuroplasticity in the PD brain, and induces a transition from an impaired PD brain organization towards a reparative one, characterized by a more balanced integration and segregation of information flow. These training-induced changes are related to improved motor performance and thus improved motor control. To our knowledge, this is the first time that graph theory indexes of resting-state EEG data have been used to reveal meaningful relationships to underlying PD processes and outcomes of importance in a translational inquiry. The incorporation of a fully digital version of PT in the form of computerized serious games adds to the innovations brought about by our study.

## Materials and Methods

### Subjects

This is a study involving 14 PwPD (male: 12, mean age: 65.5±7.12, years of education: 13.64±3) on stage II or III according to the unified Parkinson’s disease rating scale (UPDRS) (11 subjects on stage II, mean UPDRS score: 24±6.04). The subjects underwent PT, as well as psychosomatic evaluation before and after the intervention. This study is part of the pdLLM clinical trial, registered with ClinicalTrials.gov (identifier code NCT04426903). The study’s protocol was approved by the Bioethics Committee of the Medical School of the Aristotle University of Thessaloniki and was conducted under the Helsinki Declaration of Human Rights. All participants signed written informed consent before their inclusion in the study.

### Intervention Protocol

The intervention consisted of PT. The study protocol was developed to improve the quality of life of people at risk for neurodegeneration such as PwPD. The training sessions were computerized, center-based, and conducted under supervision. The sequence of training methods was pseudo-randomized and counterbalanced. The details of the PT intervention have been previously described in detail [62], [63] (summarized here in Figure 4). In short, PT is fully computerized here through the WebFitForAll facility, a web-service system through a general-purpose interface [64] and specific (serious) game-based activities of balance, endurance, flexibility, and aerobic exercise. The software has been used in many other studies (e.g., [29], [65]) with beneficial results. The intervention lasted 10 weeks, with a frequency of two times per week for an hourly session, and took place at the Thessaloniki Active and Healthy Ageing Living Lab (Thess-AHALL) [66] and the Association of Parkinson’s Patients and Friends of Northern Greece.

**Figure 4.**
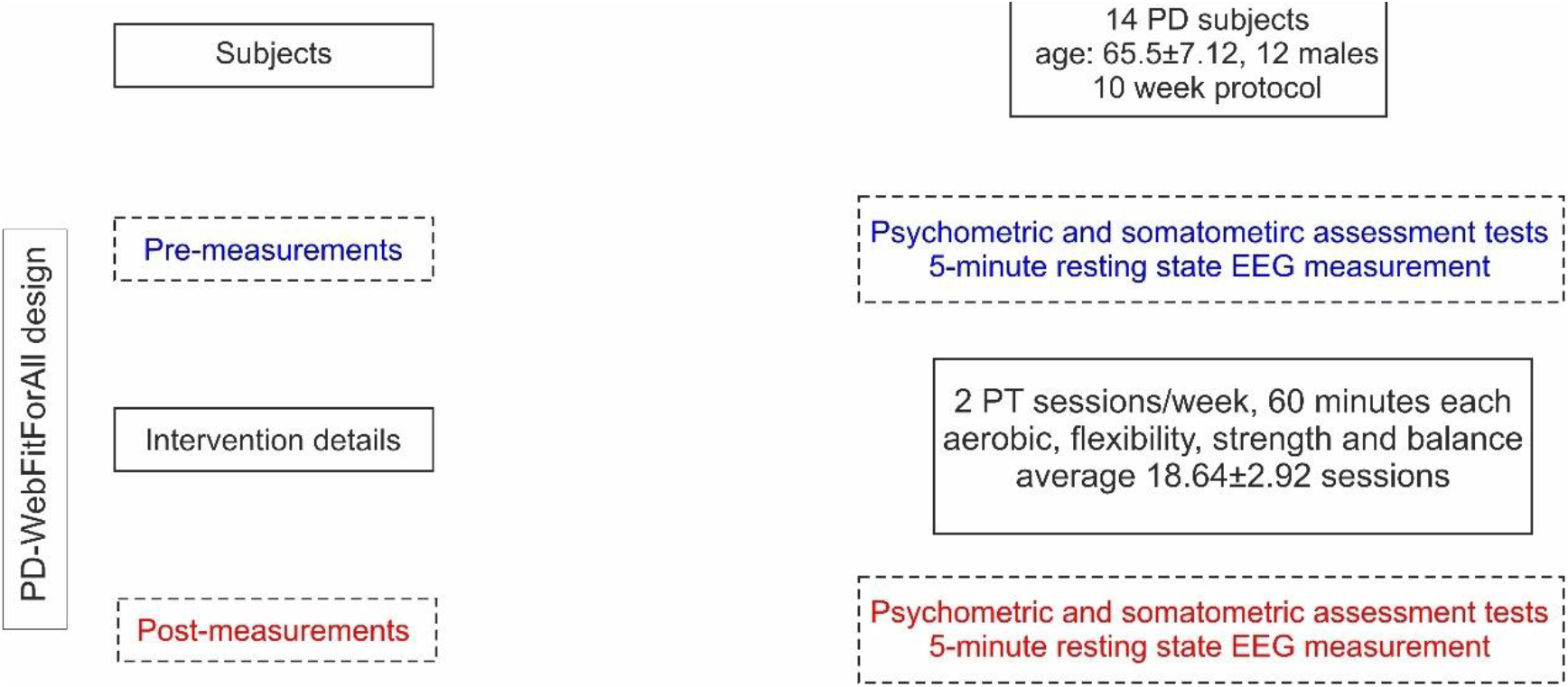
PD-WebFitForAll design and flow of PD participants

### Physical training

PT is based on the WebFitForAll exergaming platform [64], which employs motion sensor devices (i.e., Kinect), allowing the users to practice and maintain their physical status and well-being. The training protocol consists of aerobic (cycling, in-place-hiking), flexibility (stretching), strength (weightlifting and resistance), and balance (static and dynamic) exercises in compliance to the ACSM and AHA recommendations [67]. Sports experts / physical educators and physiotherapists have defined a 50-70% intensity level of the maximum heart rate (HRmax). The 10-minute warm-up phase precedes the main body of training (40 minutes) that is followed by 5 minutes’ full recovery. For the aerobic exercises, the users are transferred to a virtual environment through Google maps and explore cities or landscapes. Upon correct completion of the flexibility and strength exercises, the trainees were progressively rewarded with an array of pleasing images. Balance exercises required the movement of the body in a horizontal or vertical axis [68].

### Psychometric and somatometric assessments

The participants underwent assessments of their cognitive and physical capacity. The psychometric evaluation assessed attention, memory, and executive function, verbal fluency, mental flexibility, processing speed, and depression or well-being (MMSE [46], MoCA [47], Trail Making Test [69], PDQ-8 [48], Geriatric Depression Scale (GDS) [70]) The somatometric evaluation assessed the overall physical status, balance, walking, and risk of falls, via the 10 Meter Walk Test [45], the Community Balance & Mobility Scale [42], the Short Physical Performance Battery [41], the Fullerton Senior Fitness Test [43], the Berg Balance Scale [40], and the Tinetti Test [44].

### Experimental design and EEG recordings

Pre- and post-intervention EEG resting-state activity was recorded measured for 5 minutes, using a high-density Nihon-Kohden EEG device (128 active scalp electrode) at a sampling rate of 1000Hz. The EEG recordings were performed in an electrically shielded, sound, and light attenuated booth. The electrode impedances were kept lower than 10 kΩ. Participants were instructed to keep their eyes closed and to maintain a resting yet wakeful condition.

### EEG analysis

#### Preprocessing

The raw EEG data were initially visually inspected, bad channels were interpolated, and ocular artifacts (blinks and horizontal movement) were corrected through adaptive artifact correction [71] using the Brain Electrical Source Analysis software (BESA research, version 6, Megis Software, Heidelberg, Germany) (Figure 5, orange section). The pruned data were imported in the Fieldtrip Matlab toolbox [72] for further processing. The EEG data were filtered using a high-pass IIR filter at 1 Hz, a notch IIR filter at 47-53 Hz, and a low-pass IIR filter at 97 Hz. Independent component analysis (ICA) [73] was applied to the filtered EEG data, and the artifactual components were rejected. The EEG data were further inspected, and all remaining visible artifacts were removed. We randomly selected 15 segments, each with a duration of 4 seconds, from each EEG recording for further processing.

**Figure 5.**
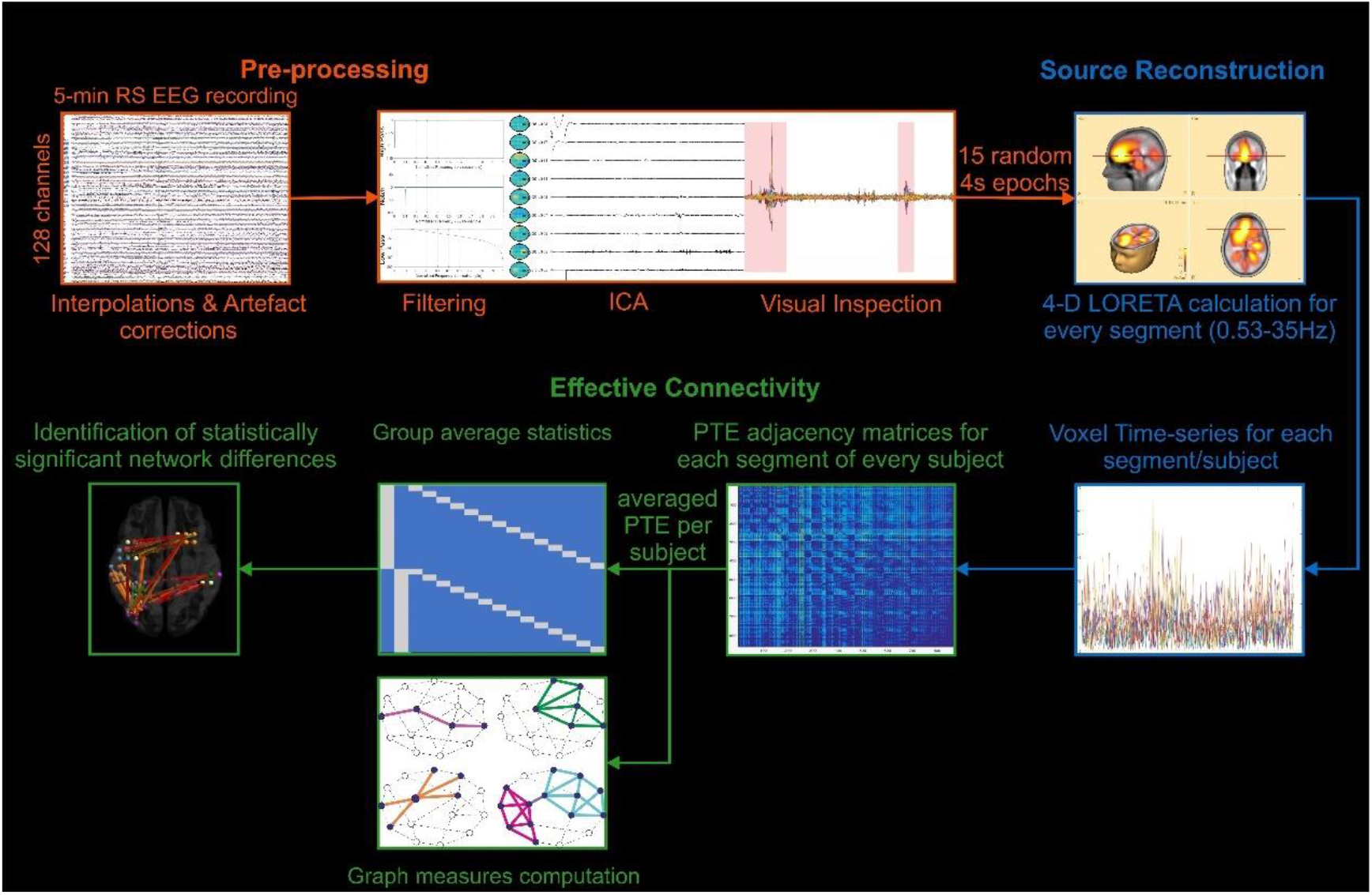
EEG data analysis schematic. **Pre-processing** (orange): EEG data were first high-pass, bandpass, and low-pass filtered. ICA and visual inspection were used to reject artefactual data. Following, 15 segments of 4-s were randomly selected. **Source reconstruction** (blue): The data were frequency filtered within 1-30 Hz, source reconstructed (4-D LORETA), and an 863-node homemade atlas was used to extract the time-series of every voxel from every segment per subject. **Functional connectivity** (green): Directed functional connectivity was computed for every segment of every subject, using the phase transfer entropy metric (PTE), and the 15 matrices of every subject were averaged into one. A network science approach was taken for the computation of graph measures per subject. Group average statistics were calculated to identify the statistically significant differences of post- and pre-intervention networks.

#### Source reconstruction

Each subject’s segments (15 segments, 4-sec duration each) were imported into BESA (Figure 5, blue section). The current density reconstructions (CDR) were estimated in the 1-30 Hz frequency band for every sample point, solving the inverse problem with the use of the LORETA [38], which does not require the *a priori* declaration of the number of sources and is suitable for whole cortex analysis. The CDRs were exported as four-dimensional images (4-D), which were, in turn, imported into Matlab. The imported images were further processed by superimposing a cortex mask including only grey matter, which excludes the brainstem and cerebellum, to limit the source space [74]. The source space consisted of 863 voxels.

#### Functional connectivity

The exported time-series of each voxel from the 4-D images were used to compute the Phase Transfer Entropy (PTE) (Figure 5, green section) [39]. Each voxel was considered as a node of the calculated network. The computation resulted in 863×863 adjacency matrices, with the metric calculated independently for every voxel of every segment. The benefit of using PTE is that its results are not based on a specific data model because its computation relies on non-linear probability distributions. This allows for the detection of higher-order relations in the phase information flow and renders the measure impervious to source leakage [39]. The adjacency matrices, which were calculated separately for every segment, were averaged, resulting in a single connectivity matrix per subject. This methodology was previously used for a cross-sectional MEG resting-state analysis [75].

### Graph measures computation

The Brain Connectivity Toolbox [76] was used to compute the graph measures of global (transitivity) and local clustering coefficient, global and local efficiency, and node betweenness centrality for each participant. The node degree centrality was also computed with the use of a Matlab function. The density of each graph was calculated by summarizing all the weights of the graph.

CPL is the average shortest path length of the edges connecting the nodes of the network [77], and GE is the average inverse shortest path length of the network [78]. TS of a graph is the ratio of closed triplets to the maximum number of triplets (open and closed) [79]. An open triplet is three nodes with one and/or two connections between them, while a closed triplet is three nodes with three connections between them (i.e., a triangle). CC of a node is the ratio of its connected neighbors to the maximum number of possible connections [77]. LE of a node is the computation of GE on a local level, strongly related to CC [15]. BC measures the importance of a node in the communication of the network’s other nodes and corresponds to the fraction of all shortest path that passes through the node [76]. DC identifies the number of connections inward and outward of a node, i.e., it indicates that the higher the degree, the more prominent a node is to the network.

Possible integration shifts were examined through CPL and GE, which quantify the efficacy of information transference and its assimilation in the network [76]. TS and CC were computed to investigate the training’s effect on robustness, and to determine if the intervention could affect segregation through the clustering organization of the PD brain network [76]. Similarly, LE was used to interpret any changes in the segregation of the network. BC quantifies the importance of a node in the information flow between nodes [76].

#### Statistical analysis

The pre- and post-intervention somatometric and psychometric assessment scores were compared using non-parametric Wilcoxon tests and paired t-tests. The statistical comparisons were performed using the IBM SPSS 25.0 software. Wilcoxon tests were performed on the psychometric battery tests and somatometric tests with a discrete-values score, while paired t-tests were applied on the remaining somatometric assessment tests.

The Network Based Statistics (NBS) [80] toolbox was employed to estimate the statistically significant differences between the pre- and post-intervention connectivity networks. A paired-samples t-test was performed, with the significance threshold defined at *p<0*.*05*, corrected using 10000 random comparisons via False Discovery Rate (FDR) correction. The significant differences between the two time points were visualized as weighted graphs through the BrainNet Viewer [81] toolbox. DC was calculated from the outcome of this comparison, and the results were depicted in the same graph.

Analysis of Covariance (ANCOVA) was used for the graph measures. Each measure was the dependent variable with density being the covariate because density seriously affects the computation of the other measures. For local measures (i.e., CC and BC), ANCOVA with FDR correction on the p-values for Type I errors was performed for each of the 863 nodes.

The relationship between graph measures and assessment scores was examined by employing the linear regression model, which can determine the possible prediction of a dependent variable via an explanatory one (independent variable). The linear regression model was calculated on the difference of pre- and post-training measurements with elements of both graph measures and assessment scores set as the independent variable. Thus, we investigated whether shifts in the cortical organization could predict the beneficial impact of the intervention on the physical and cognitive capacity of our subjects and the opposite. The statistical significance of the prediction was tested by F-test at a significance level of *p < 0*.*05*.

## Data Availability

The datasets generated during and/or analysed during the current study are available from
the corresponding author on reasonable request.

## Competing interests

There are potential conflicts of interest (other, not financial, outside the scope of the submitted work) for the author P. Bamidis in respect of the Aristotle University of Thessaloniki (AUTH). WebFirForAll has been developed at AUTH, as an extension of FitForAll (FFA), originally developed in AUTH during the Long Lasting Memories (LLM Project) (www.longlastingmemories.eu), funded in the beginning by the ICT-CIP-PSP Program of the European Commission. It now forms part of LLM Care (www.llmcare.gr/en), a technology transfer/self-funded initiative that emerged as the non-for-profit business exploitation of LLM.

## Acknowledgments

This work was partly supported by the EU H2020 PHC 2014 2015/H2020 PHC 2015, grant agreement N 690494: ‘i Prognosis’ project (www.iprognosis.eu) and partly supported by the European CIP ICT PSP.2008.1.4 Long Lasting memories (LLM) project (Project No. 238904) www.longlastingmemories.eu), as well as, its subsequent business exploitation scheme, namely, LLM Care, which is a self-funded initiative at the Aristotle University of Thessaloniki (www.llmcare.gr). The study has also benefited from an internship program for undergraduate students, funded by the European Social Fund and co-financed by Greek National Resources through the Operational Program “Competitiveness, Entrepreneurship and Innovation” of the Corporate Pact for the Growth Framework 2014-2020, through the Intermediate Managing Agency on “Human Resources, Education and Lifelong Learning” of the Greek Ministry. The authors would like to thank all the participants, their families, and caregivers, the Association of Parkinson’s Patients and Friends of Northern Greece, as well as Fotini Dolianiti, Antonios Gantaras, Stylianos Gavriilidis, Sotiria Gylou, Georgina Ilia, Katerina Katsouli, Maria Meitanidou, Ioannis Papadopoulos, Tzortzina Papaioannou, and Dimitrios Sarris (in alphabetical order) for pilot execution. The authors would also like to acknowledge the contribution of technical support provided by Theodoros Savvidis, Athina Grammatikopoulou, and Michael Stadtschnitzer. Clinical expertise has been provided by Neurologists Sevatsi-Maria Bostanjopoulou and Zoe Katsarou who are also the scientific advisors of the Association of Parkinson’s Patients and Friends of Northern Greece. In addition, the authors would like to acknowledge the support of the iPrognosis consortium, led by Prof. Leontios Hadjileontiadis.

## Notes

### Clinical Trial

NCT04426903

### Funding Statement

No external funding was received

### Author Declarations

Bioethics Committee of the Medical School of Aristotle University of Thessaloniki

